# Study protocol for development of an options assessment toolkit (OAT) for National Malaria Programs in Asia Pacific to determine best combinations of vivax radical cure for their given contexts

**DOI:** 10.1101/2022.07.22.22277913

**Authors:** Manash Shrestha, Josselyn Neukom, Sanjaya Acharya, Muhammad Naeem Habib, Lyndes Wini, Tran Thanh Duong, Ngo Duc Thang, Karma Lhazeen, Kamala Thriemer, Caroline A. Lynch

## Abstract

**Introduction:** Recent advances in G6PD deficiency screening and treatment are rapidly changing the landscape of radical cure of vivax malaria available for National Malaria Programs (NMPs). While NMPs await the WHO’s global policy guidance on these advances, they will also need to consider different contextual factors related to the vivax burden, health system capacity, and resources available to support changes to their policies and practices. Therefore, we aim to develop an Options Assessment Toolkit (OAT) that enables NMPs to systematically determine optimal radical cure options for their given environments and potentially reduce decision-making delays. This protocol outlines the OAT development process.

**Methods:** Utilizing participatory research methods, the OAT will be developed in four phases where the NMPs and experts will have active roles in designing the research process and the toolkit. In the first phase, an essential list of epidemiological, health system, and political & economic factors will be identified. In the second phase, 2-3 NMPs will be consulted to determine the relative priority and measurability of these factors. These factors and their threshold criteria will be validated with experts using a modified e-Delphi approach. In addition, 4-5 scenarios representing country contexts in the Asia Pacific region will be developed to obtain the expert-recommended radical cure options for each scenario. In the third phase, additional components of OAT, such as policy evaluation criteria, latest information on new radical cure options, and others, will be finalized. The OAT will be pilot-tested with other Asia Pacific NMPs in the final phase.

**Ethics and Dissemination:** Human Research Ethics Committee approval has been received from the Northern Territory, Department of Health, and Menzies School of Health Research (HREC Reference Number: 2022-4245). The OAT will be made available for the NMPs, introduced at the APMEN Vivax Working Group annual meeting, and reported in international journals.

**Strengths and limitations of the study:**

▸ We will systematically develop the OAT in collaboration with select NMPs who represent the intended end-users of the toolkit, thus ensuring this research project is relevant to NMP decision-making on vivax radical cure treatment and approaches.
▸ This study will obtain expert consensus on radical cure options for particular scenarios and threshold criteria for baseline factors.
▸ While 4-5 scenarios will be developed in the OAT, it is possible that some individual country contexts of the Asia Pacific region may not be fully represented in those scenarios.
▸ Some larger malaria endemic countries in the Asia Pacific have a wide diversity of contexts at the sub-national level when it comes to vivax malaria. However, as the policies are made at the national level, the OAT will not be designed to facilitate analysis of separate radical cure options for the sub-national level.
▸ The toolkit may be subject to change/revision as new data on the radical cure tools become available and as additional countries use the OAT post the APMEN Vivax Working Group annual meeting.

## INTRODUCTION

Effective radical cure of the dormant liver forms of *Plasmodium vivax*, preventing relapses, is critical for the elimination of malaria. As vivax endemic countries intensify their malaria control efforts, malaria decreases, however often *P. vivax* becomes the dominant parasite.^1^ A recent analysis suggests that more than 85% of acute episodes are caused by relapses and this proportion might even be higher in some regions e.g. the Greater Mekong Subregion.^2 3^

Currently, the World Health Organization’s (WHO’s) recommended radical cure is low dose primaquine (PQ) given for 14 days.^4^ This relatively long treatment regimen is challenging and often results in poor health worker compliance and patient adherence.^5 6^ In addition, until recently, limited point-of-care (PoC) diagnostic options were available to identify patients with glucose-6-dehydrogenase (G6PD) deficiency at risk of drug-induced haemolysis. Recent advances in the tools available to tackle *P. vivax* relapses, including shorter and higher dose treatment regimens with primaquine (PQ),^7^ single dose tafenoquine (TQ),^8 9^ and novel quantitative PoC G6PD tests (SD biosensor) are changing the landscape of tools becoming available to national malaria programs (NMPs).^10 11^ Global policy recommendations addressing the use of new tools are expected from the WHO’s Global Malaria Program but will need to be adapted to country contexts.

As a result, NMPs are faced with determining which, among these options, are best for their given contexts while accounting for their vivax and G6PD epidemiology, health system capacity, and political and economic factors. Ruwanpura et al. (2021) demonstrated that policy change processes in the Asia Pacific region are nebulous and opaque, taking up to three years in some cases to move from evidence availability to policy change.^12^ Given most countries in the region aim to eliminate malaria by 2030, the possibility of doing that could be constrained by slow decision-making processes. Research has shown that having multiple options for decision-makers to choose among can delay decision-making (e.g. in HIV).^13^ A toolkit that enables NMPs to systematically determine which radical cure options are best for their given environments, has the potential to reduce decision-making delays and accelerate availability of new tools, thus reducing morbidity and mortality related to vivax malaria.

Additionally, different NMPs in the Asia Pacific have identified needing support to decide the optimal combination of vivax radical cure options as a priority in the coming 1-2 years as different radical cure advances become available.^14^ As such, this research and tool development responds to an immediate need identified by NMPs in the region. Irrespective, several similar toolkits exist for other diseases and medical areas that have facilitated decision making by disease programs and stakeholders in the past.^15^ However, reviews of public health toolkits reveal that only around 10% of the toolkits are designed for health decision makers/policy makers and the uptake of toolkits can vary highly.^16 17^ In addition, there are limited availability of specific details of toolkit development methods, especially for malaria.

Therefore, the aim of this work is to develop and pilot an options assessment toolkit (OAT) that enables national decision-makers determine the optimal set of tools for better radical cure of vivax malaria. While the NMPs await WHO’s global recommendations, the OAT seeks to provide evidence-informed support to boost NMP’s technical capacity and assist in nationally-owned decision making.

## METHODS AND ANALYSIS

### Study design

The OAT will be developed through participatory research methods in a sequential multi-phase design (Figure 1), where NMPs and regional experts will be engaged and have active roles in the research process and jointly contribute to the OAT development. Using this method, we aim to ensure the research is relevant to NMP decision-making on vivax radical cure treatment and approaches.^18^ The foundational premise of participatory research methods is the value placed on genuine and meaningful participation – thus, any meetings and discussions held with NMPs and experts will include adapted in-depth interviews (IDI), participatory group discussions, and Delphi process. The aim in adapting these approaches is to ensure that engagement with NMPs and regional experts empowers their “ability to speak up, to participate, to experience oneself and be experienced as a person with the right to express yourself and to have the expression valued by others”.^19^

**Figure 1.**
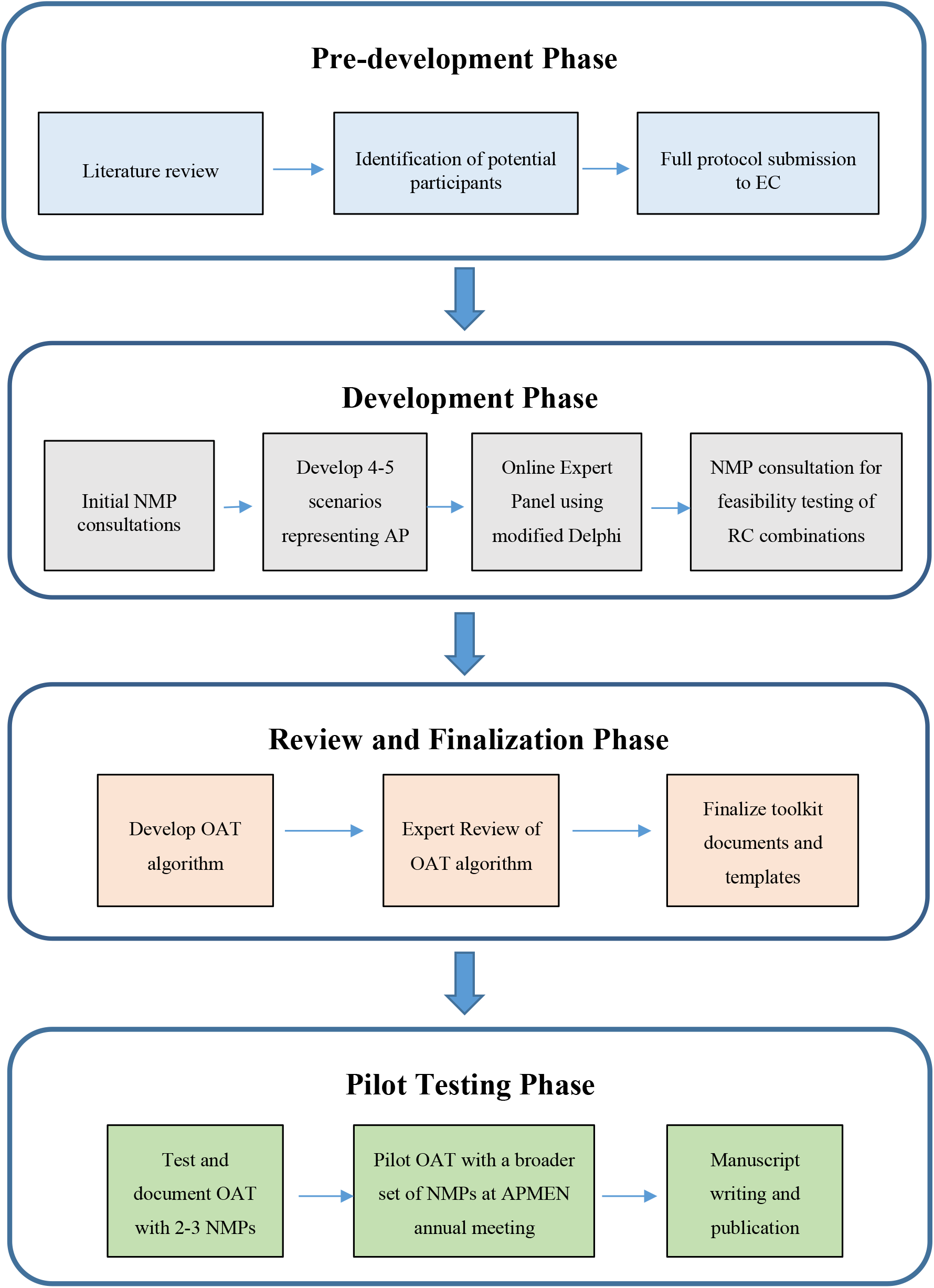
OAT development study design.

### Study population

NMPs in the Asia Pacific region are the intended users of the OAT and co-developers of the tool. NMPs are the national level governmental officers in the respective Ministries of Health who are designated for malaria control and responses in the country.

Along with the NMPs, the toolkit development process will also include consultations (e.g. Delphi process) with global/regional experts. For operational feasibility, experts will be defined as international or regionally recognized individuals with more than 10 years of professional experience and high-level expertise in malaria.

Methodologists who have experience of developing similar public health toolkits will also be consulted separately to strengthen the methodology of OAT development.

The participants in this toolkit development can be classified as shown in Table 1.

**Table 1.**
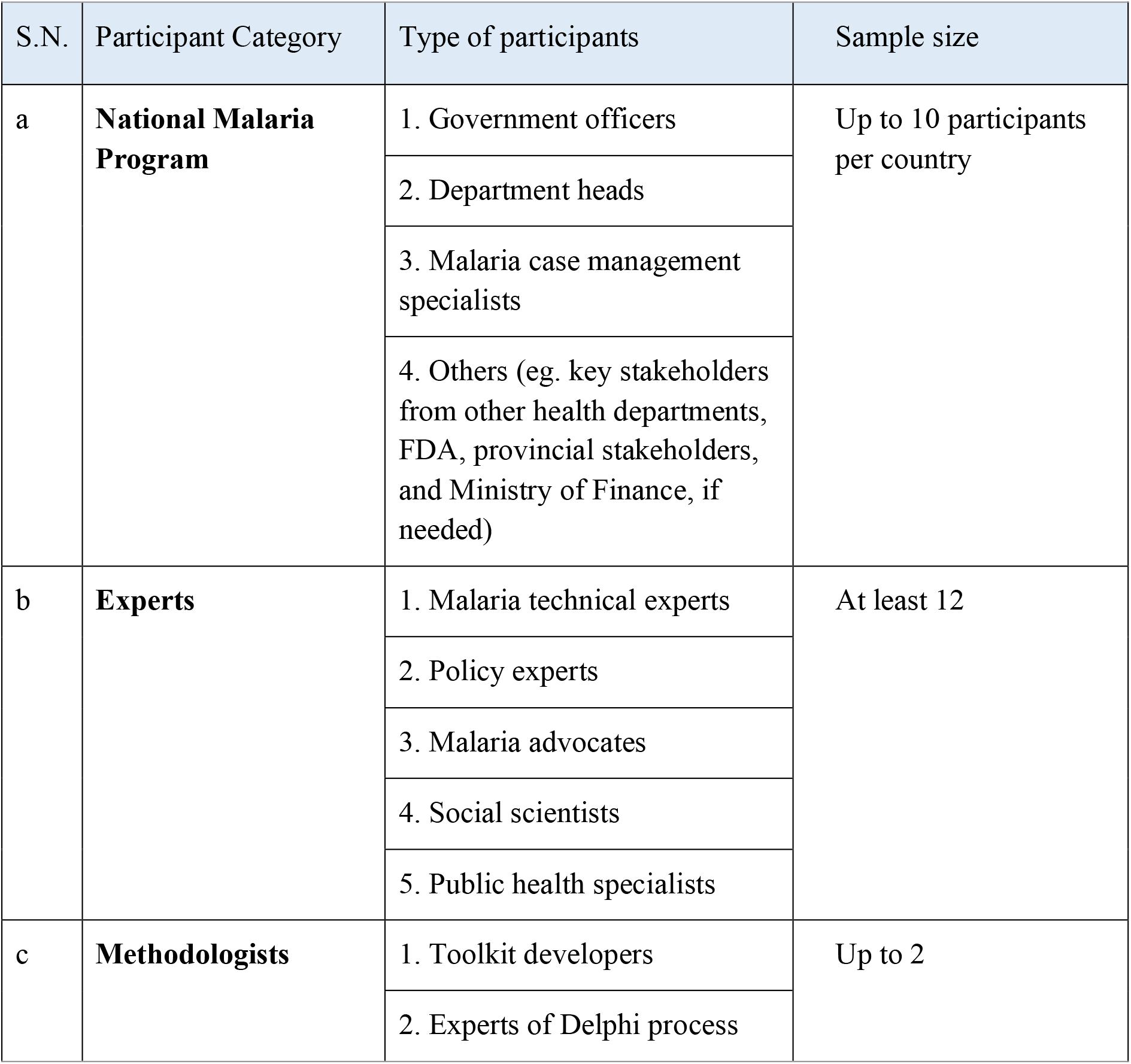
Participants in OAT development process.

#### Participant selection

A selection process was undertaken to identify 2-3 potential NMPs from different parts of the Asia Pacific region to approach for inviting to the OAT development process using the criteria outlined in Table 2.

**Table 2.**
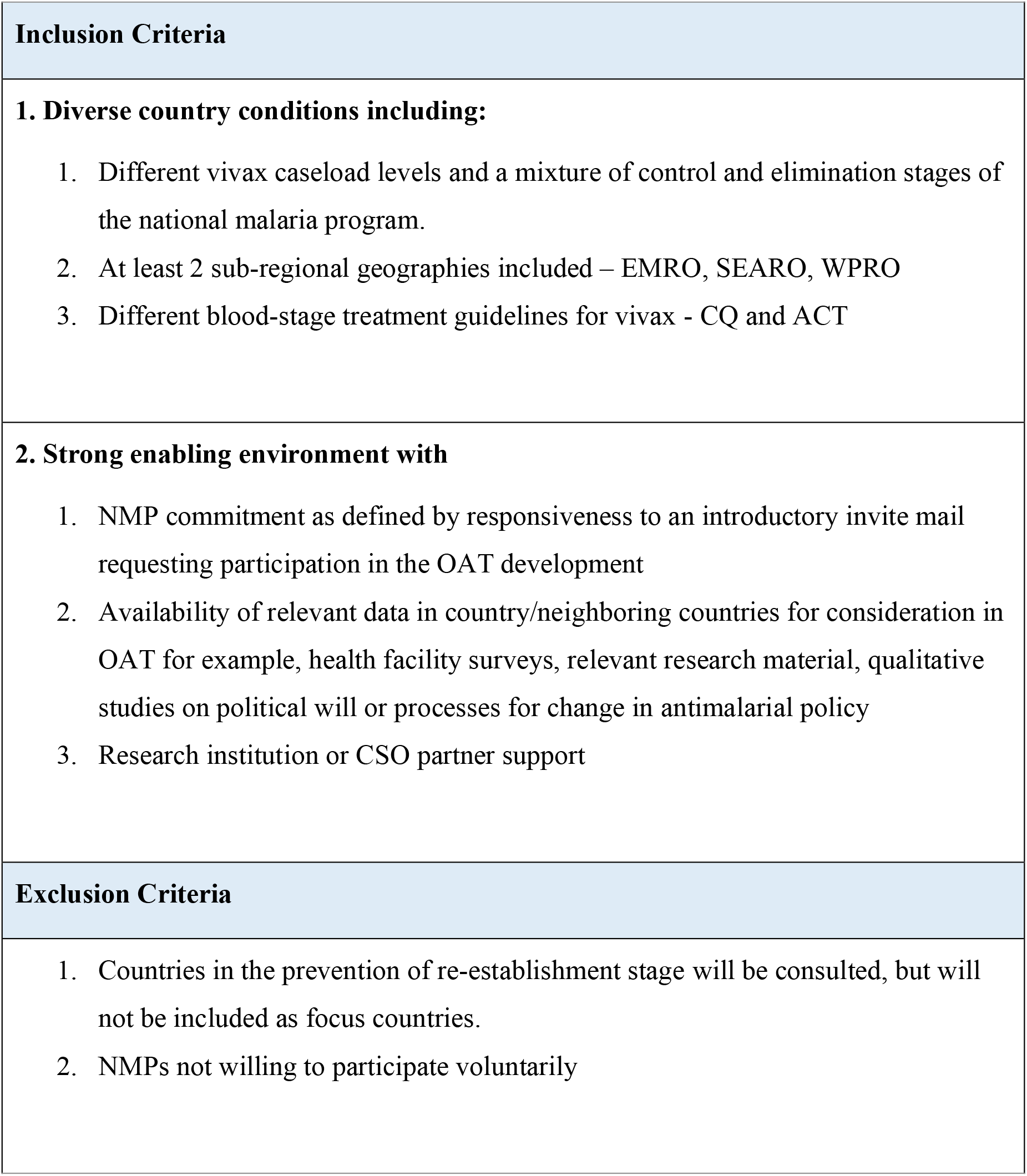
NMP country selection criteria.

| **Inclusion Criteria** |
| --- |
| **1. Diverse country conditions including:**<br>1. Different vivax caseload levels and a mixture of control and elimination stages of the national malaria program.<br>2. At least 2 sub-regional geographies included – EMRO, SEARO, WPRO<br>3. Different blood-stage treatment guidelines for vivax - CQ and ACT |
| **2. Strong enabling environment with**<br>1. NMP commitment as defined by responsiveness to an introductory invite mail requesting participation in the OAT development<br>2. Availability of relevant data in country/neighboring countries for consideration in OAT for example, health facility surveys, relevant research material, qualitative studies on political will or processes for change in antimalarial policy<br>3. Research institution or CSO partner support |
| **Exclusion Criteria** |
| 1. Countries in the prevention of re-establishment stage will be consulted, but will not be included as focus countries.<br>2. NMPs not willing to participate voluntarily |

The experts will be selected using the criteria as shown in Table 3.

**Table 3.**
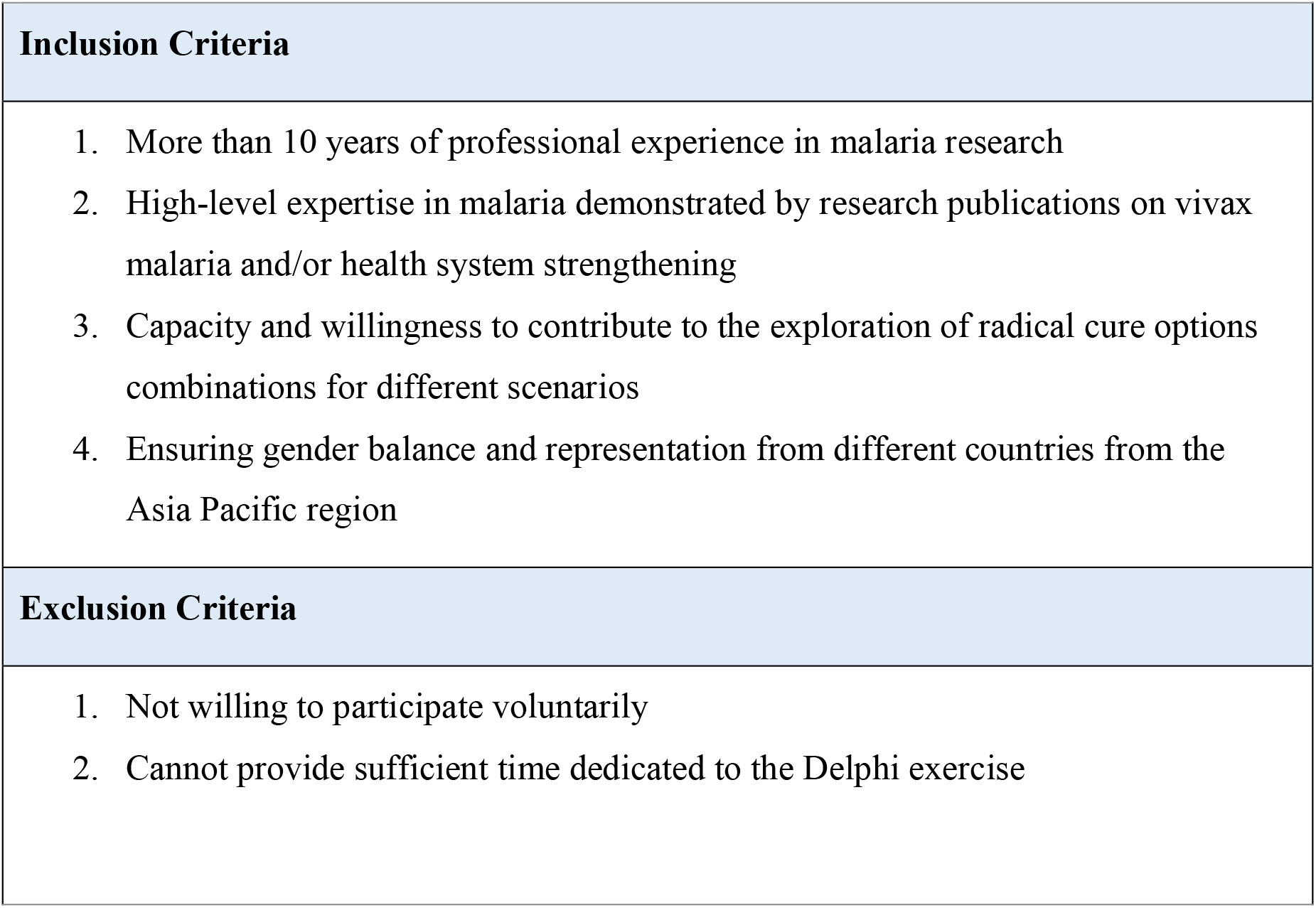
Expert selection criteria.

### Study Phases

#### 1. Pre-development phase

The definition of “toolkit” was adapted from Barac et al., as “the package of multiple resources that codify explicit knowledge, such as templates, guidelines, algorithms, summaries, and that are geared to knowledge sharing and/or facilitate change in policy and practice”. ^16^ In this preliminary phase, a literature review was conducted that scoped out the systematic reviews of public health toolkits and similar toolkits developed for health policy makers. The review indicated that the evidence base on health toolkits is limited, with only around 10% of the toolkits being designed for health decision makers/policy makers.^16^ Most of the toolkits did not have their contents well-described and only two-thirds of the toolkits (26/39) specifically indicated the clinical evidence, rationale or theoretical basis underlying the toolkit strategies.^16 20^ While the users of existing public health toolkits were highly satisfied with the toolkit, usefulness of individual tools varied and so did the uptake of these toolkits.^16^ A further review of toolkits similar to the expected OAT revealed limited availability of specific details of toolkit development methods, especially for malaria (Supplementary Table 1). Nonetheless, learning from the processes of previous toolkit development has helped inform OAT development methodology,^15^ with a keen focus on ensuring that the OAT is useful for decision-makers. Additionally, a list of key epidemiological, health system, and political & economic variables was identified for consideration to include as variables in consultation with NMPs (Table 4).

**Table 4.**
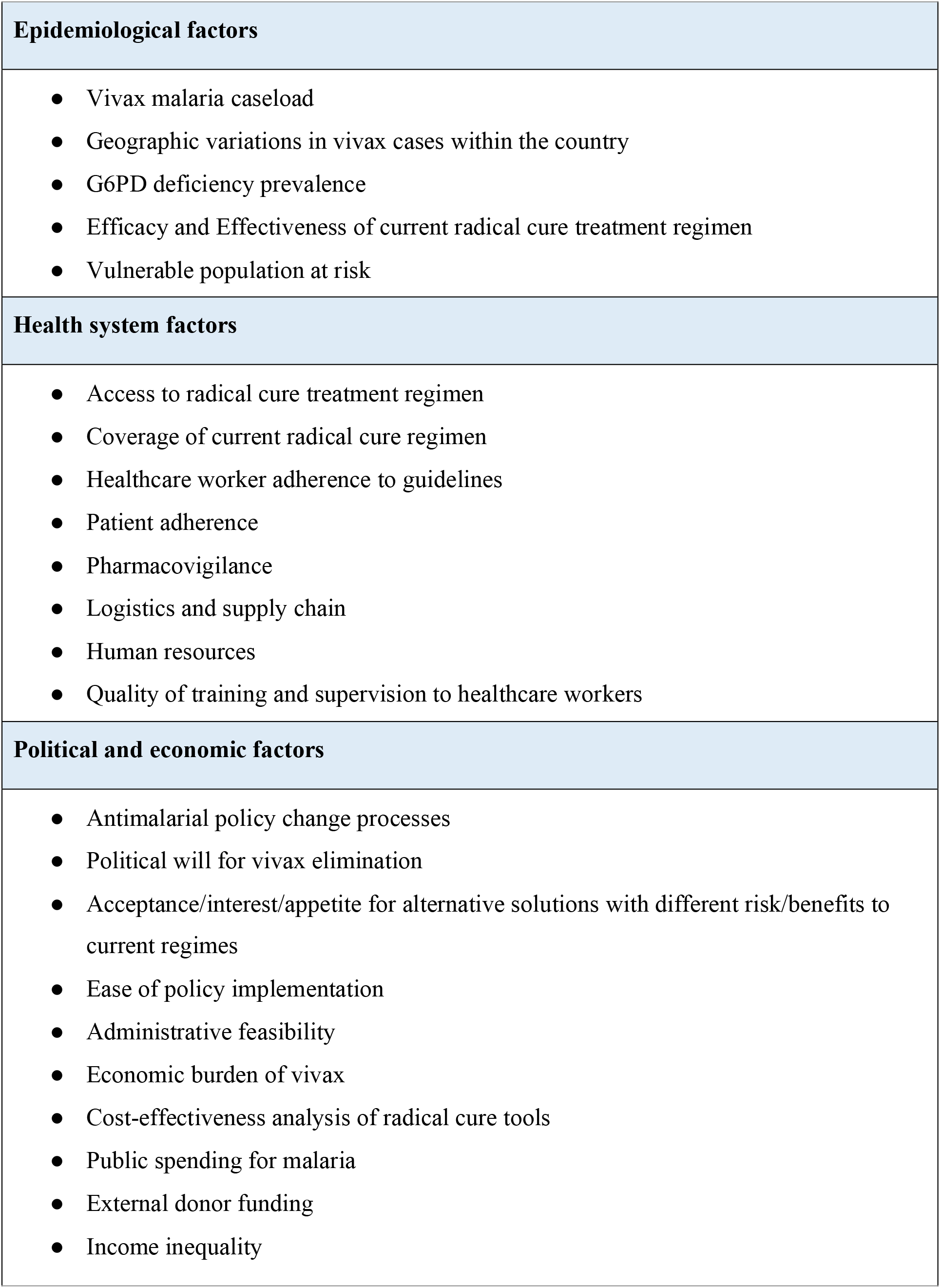
Initial list of key variables for discussion with NMPs.

| **Epidemiological factors** |
| --- |
| • Vivax malaria caseload<br>• Geographic variations in vivax cases within the country<br>• G6PD deficiency prevalence<br>• Efficacy and Effectiveness of current radical cure treatment regimen<br>• Vulnerable population at risk |
| **Health system factors** |
| • Access to radical cure treatment regimen<br>• Coverage of current radical cure regimen<br>• Healthcare worker adherence to guidelines<br>• Patient adherence<br>• Pharmacovigilance<br>• Logistics and supply chain<br>• Human resources<br>• Quality of training and supervision to healthcare workers |
| **Political and economic factors** |
| • Antimalarial policy change processes<br>• Political will for vivax elimination<br>• Acceptance/interest/appetite for alternative solutions with different risk/benefits to current regimes<br>• Ease of policy implementation<br>• Administrative feasibility<br>• Economic burden of vivax<br>• Cost-effectiveness analysis of radical cure tools<br>• Public spending for malaria<br>• External donor funding<br>• Income inequality |

#### 2. Development Phase

The first set of tools of OAT will be developed in this phase. Methodologists (n=1-2) will be consulted in this phase to refine the OAT methodology as needed. Availability of relevant national data as pre-identified in Table 4 will be explored from the NMPs, and an expert panel will be consulted to determine optimal combinations of available radical cure tools for various scenarios of Asia Pacific region. This phase will include the following steps:

##### Development of readiness assessment/situational analysis template

The list of initial variables (Table 4) identified in the pre-development phase will be explored further by collating national literature, particularly the gray literature found in national reports, project data, monitoring and evaluation reports. Using the variables and national-level information, a template for assessing the readiness of the national program to improve the radical cure of vivax malaria will be developed for NMPs. This template will be aided by guidance on how to source relevant data.

##### Initial phase of NMP consultations

The selected NMPs will be consulted to determine measurability and priority of the key variables identified. The consultation will be conducted either through face-to-face meetings or in the form of online in-depth interviews using interview guides, considering data and literature available on national contextual factors. Other stakeholders such as key partners, drug authorities, provincial programs etc., may also be included in the consultations if identified by the NMPs.

Where necessary, the in-country policy change process for malaria will be mapped and any key variable or evidence that is required for that process that should be included into the OAT will be identified. The outputs from the initial consultations will be used to draft a list of prioritized and measurable variables that should be considered when deciding on optimal vivax radial cure per scenario.

##### Development of scenarios representing Asia Pacific region

After the NMP consultations, the findings will be used, in addition to available literature, to develop 4-5 scenarios representing different epidemiological, health system and political economic contexts for the Asia-Pacific region. The developed scenarios will be presented to the NMPs for their feedback with a particular focus on how well they can identify with the scenarios and whether there are any significant gaps. Revised scenarios will then be presented to the experts to match optimal radical cure tools for each scenario. Different contextual and health system scenarios will be beneficial for the NMPs to see what the experts think and visualize potential future scenarios as burden reduces in higher-burden countries. The different scenarios might also present an opportunity to assess resource requirements to support the use of different tools for each scenario and flag additional investments required to facilitate certain options e.g. strengthening capacity at peripheral levels, increasing budget allocation, etc.

##### Expert panel consultation

A list of regional/global experts will be identified and approached for an online expert panel consultation. ^21^ Using a modified e-Delphi process,^22^ multiple rounds of consultations will be conducted with the experts to validate the factors in readiness assessment template, determine threshold criteria for these factors, and match optimal radical cure combinations to regional scenarios. While Delphi sample sizes depend more on group dynamics in reaching consensus than their statistical power, a minimum of 12 respondents is generally considered to be sufficient.^23^ Therefore, we will invite 25 experts to the Delphi process to enroll at least 12, assuming the response rate to be around 50%. A detailed methodology for the Delphi is provided in Supplementary File 1.

##### Second phase of NMP consultations

The NMPs will be followed-up after the initial consultations to clarify their priorities and explore key factors for decision-making. Again, after receiving agreed scenario-matched optimal radical cure combinations from the experts, another round of consultations will be held with the NMPs to review and test the feasibility of the radical cure combinations provided by the experts. Any concerns or issues raised by the NMP that could lead to decision making delays for radical cure will be documented.

#### 3. Review and Finalization Phase

In this phase, the final set of tools of the OAT will be developed as described below:

##### OAT algorithm

Based on the discussions with NMP and experts, an OAT algorithm or decision tree will be constructed that will include different criteria for consideration for use with NMPs. The OAT algorithm will be reviewed by the experts before using it with NMPs. The experts will be consulted virtually for their comments on the OAT algorithm and if necessary, a virtual round-table discussion session will be held afterwards.

##### Other tools

Other tools to support the OAT algorithm will also be developed that can include:

- A step-by-step guidance on how to use the OAT toolkit (based on documentation of the process and engagement with NMPs)
- Accessible evidence briefs on efficacy and effectiveness of current radical cure drugs and latest information on high sensitivity rapid diagnostic tests (HS-RDTs), G6PD screening tests, and radical cure options near end of pipeline
- NMP weighting tool for different variables
- Approaches for optimized radical cure tools
- Policy options evaluation matrix
- Policy uncertainties and potential mitigation actions template

#### 4. Pilot Testing Phase

In this final phase, the final OAT developed will be first tested with the co-developing NMPs and then introduced for feedback from a broader set of NMPs from the Asia Pacific:

##### Test with co-developing NMPs

The participating NMPs will be consulted for testing the OAT. The sessions will be documented to understand NMPs perspectives on usefulness and ease of using OAT. Any modifications needed to improve the OAT will be made according to NMP feedback.

##### Pilot OAT with all NMPs

The final OAT version will be piloted among all the NMPs at the Asia Pacific Malaria Elimination Network (APMEN) Vivax Working Group face-to-face annual meeting in November 2022 (currently planned to be conducted in India). We will go beyond introducing the OAT to work with all participating countries to develop a plan to use the OAT in their country at the meeting. If needed, the OAT will be further revised and presented for use with NMPs at APMEN TechTalks/mini-workshops.

### Data Capture and Analysis

Throughout the process, interviews, meetings and discussions with NMPs and regional experts will be fully documented and transcribed for analysis. Data analysis will be done primarily with NMPs and to some extent with regional experts as part of the participative approach. Participants will read the data and synopses of various meetings and interviews, on the data collection process in advance. A facilitator will pose a question of interest. Pertinent to the tool development e.g. how important was this variable when thinking about choosing vivax radical cure tools in X area of your country’? This will be explored by participants. Individually, participants will brainstorm and write down what they see as the most significant themes. These themes will be shared with the full group and the facilitator to inform the discussion. New insights or understanding gained through this interaction will be captured and added to the data collection and learning process.

### Ethics and Dissemination

This protocol has been reviewed and approved by the Human Research Ethics Committee of the Northern Territory, Department of Health and Menzies School of Health Research (HREC Reference Number: 2022-4245).

The final toolkit developed will be made available for use for the NMPs and presented at APMEN annual meeting, various international talk programs, and conferences. The development and findings of the project will be reported in relevant international journals following the COREQ (Checklist for reporting qualitative research) guidelines. ^24^

## DISCUSSION

Toolkits can be an effective strategy for knowledge translation, ensuring the use of research evidence to inform healthcare decision-making.^16^ However, only a few health toolkits have focused on public health policymakers and even fewer toolkits describe how they have been developed. In this protocol paper, we have delineated the multiphasic, collaborative, and systematic plan of developing the OAT which aims to assist the NMPs and national policymakers in assessing their options for radical cure of vivax and influencing policy.

Our study design and participatory research methods provide the necessary scientific rigor that is often lacking in toolkit development.^16 18^ Consideration of both the technical factors (i.e., malaria epidemiology and health systems) and non-technical factors (i.e., political will, costs, and external influence), along with a policy evaluation criteria and other significant tools, make OAT a comprehensive toolkit for radical cure of vivax malaria. Furthermore, our qualitative research design allows flexibility during the iterative process of consultations with NMPs and experts. Importantly, collaborating with NMPs during the development process will help in the uptake and ownership of the final toolkit. In addition, findings from the Delphi process in this study will provide expert consensus on radical cure options for particular scenarios and threshold criteria for key factors such as efficacy of radical cure drugs, which has remained contentious until now.

Linking evidence into policy is seldom a linear process. This process often gets undermined and mediated by personal values, ideologies, economic interests, and organizational practices.^25^ Through our approach, where we bring researchers/experts and policy/program managers together and collaborate in this toolkit development, we hope to promote knowledge sharing and mutual understanding. Our approach also seeks to add value for the policy makers and malaria program managers who are the end users of the OAT through consultative dialogue and coproduction of insights.^25^

Despite the strengths of this study, there are a few limitations in the toolkit design. First, while 4-5 scenarios will be developed in the OAT, it is possible that some individual country contexts of the Asia Pacific region may not be fully represented in those scenarios. Second, some larger malaria endemic countries in the Asia Pacific have a wide diversity of contexts at the sub-national level when it comes to vivax malaria. However, as the policies are made at the national level, separate radical cure options for the sub-national level may not be developed explicitly in the OAT. Third, the toolkit may be subject to change/revision as new data on the radical cure tools become available. Finally, it can be challenging for the NMPs to use the toolkit if it is too complex as they may not have enough time. This can be mitigated by applying the steps of the toolkit over a certain time, as opposed to one sitting and using the APMEN TechTalks and annual meeting to simplify the toolkit.

In conclusion, this paper outlines the process of OAT development in detail to inform relevant stakeholders and assist other researchers developing similar toolkit/s in other settings for malaria or for other diseases.

## Data Availability

All data/tools that will be produced in the present study will be made available online at www.vivaxmalaria.org.

## Authors’ contributions

All authors contributed to the overall study design and specific methodologies. All authors approved the final version for the submission.

## Funding statement

This work is supported by funding from Medicines for Malaria Venture (MMV) & the Asia Pacific Malaria Elimination Network (APMEN).

## Competing interests statement

The authors declare that they have no conflict of interests.

## Notes

### Competing Interest Statement

The authors have declared no competing interest.

### Author Declarations

Human Research Ethics Committee approval has been received from the Northern Territory, Department of Health, and Menzies School of Health Research (HREC Reference Number: 2022-4245).

