## Supplementary File 1 for "Study protocol for development of an options assessment toolkit (OAT) for National Malaria Programs in Asia Pacific to determine best combinations of vivax radical cure for their given contexts"

**Detailed Methodology for Delphi in OAT development**

### Background:

Consensus or group judgment methods are used to obtain opinions from a group of people with expertise in a particular area and can be used for predicting future patterns, determining priorities, generating ideas, and solving problems. One of such techniques is the Delphi technique, which was first developed in the 1950s by Norman Dalkey and Olaf Helmer to gain reliable expert consensus while providing anonymity and avoiding direct confrontation between experts [1].

The Delphi method is a systematic way of determining expert consensus that is useful for answering questions that are not amenable to experimental and epidemiological methods [2]. This approach is predicated on a series of ‘rounds’, where a group of experts is asked their opinions on a specific issue. The questions for every round are based partially on the findings of the previous one, allowing the study to evolve over time in response to earlier findings. After each round, the experts are provided with the results of the previous round including their own responses, allowing them to reflect on the views of others and reposition their own opinions accordingly [1]. Experts get an opportunity to consider and feedback on what they perceive to be the strengths and weaknesses of other’s responses. This framework of expert opinion rounds, with each round built on previous findings and every allowing response to be reconsidered by participants, is meant to permit a consensus view that answers the research question [3].

Modified Delphi is a group consensus strategy that systematically uses literature review, opinion of stakeholders and the judgment of experts within a field to reach agreement [4]. The modified Delphi technique is similar to the traditional Delphi in terms of procedure (i.e., a series of rounds with selected experts) and intent (i.e., to predict future events and to arrive at consensus) [5]. The major modification consists of beginning the process with a set of carefully selected items which are drawn from synthesized reviews of the literature and interviews with selected stakeholders. The primary advantages of this modification to the Delphi is that it (a) typically improves the initial round response rate, and (b) provides a solid grounding in previously developed work [5].

Similarly, e-Delphi is an online method widely used in health and social research to strengthen decision-making processes and reach consensus on developing guidelines for health services [6]. The e-Delphi method allows experts to communicate and engage online in their own time until consensus is reached. It is anonymous because neither the researcher nor the experts are physically present, which might influence the communication and lead to prejudice.

### Purpose:

A modified e-Delphi will be used during the OAT development to obtain expert consensus for the following objectives:

1. To validate the factors developed for the readiness assessment template
2. To determine threshold criteria for the decision making factors included in the readiness assessment template
3. To validate the developed scenarios of vivax elimination readiness in the Asia Pacific region
4. To obtain expert-recommended options of radical cure test and treat combinations for each scenario

### Methodology:

In the OAT development, a modified e-Delphi will be used where the first round of expert opinion wiil be preceded by identification of readiness assessment factors and scenario development which entails a literature review and consultations with select national malaria programs. Multiple rounds of expert opinions will be conducted online via a web-based questionnaire ([www.paperform.co](http://www.paperform.co)) and reported according to the guidelines on conducting and reporting of Delphi studies (CREDES)[7].

Key elements include having a facilitator who brings together the group of experts, provides questionnaires for the expert panel, summarizes results of subsequent rounds of Delphi process to the expert panel so they may respond to the summaries, have the experts revise their responses, and finally form a consensus.

#### Definition of “Expert” and Selection Criteria

For operational feasibility, experts will be defined as global or regionally recognized individuals with more than 10 years of professional experience and high-level expertise in malaria. The following selection criteria will be used for inclusion and exclusion of experts (Table 1):

Table 1. Selection criteria of Experts for Delphi:

**Inclusion Criteria**

1. More than 10 years of professional experience in malaria research
2. High-level expertise in malaria demonstrated by research publications on different domains of vivax malaria and/or health system strengthening
3. Capacity and willingness to contribute to the exploration of radical cure options combinations for different scenarios
4. Ensuring gender balance and representation from different countries from the Asia Pacific region

**Exclusion Criteria**

1. Not willing to participate voluntarily
2. Cannot provide sufficient time dedicated to the Delphi exercise

#### Sample size

A systematic review identified that a majority of Delphis included ≤25 respondents [8]. While Delphi sample sizes depend more on group dynamics in reaching consensus than their statistical power, a minimum of 12 respondents is generally considered to be sufficient [9]. Therefore, we will invite 25 experts to the Delphi process to enroll at least 12, assuming the response rate to be around 50%.

#### Recruitment procedure

The experts will be selected carefully from APMEN’s database using the selection criteria set for this study (i.e., purposive selection). The identity of the selected experts will be kept anonymous to promote honesty and reduce the ‘*halo effect’* among the experts of different profiles for extra credence [1]. The experts will be recruited in this study using the following steps:

1. Personal invitations will be sent to the 25 purposively selected experts by the Principal Investigator and Co-Prinicipal Investigator (KT and CAL) as a strategy to improve recruitment and retention of experts in the Delphi process [10].
2. After establishing a personal communication, the experts will be provided with an introductory email and information about the OAT.
3. The experts will be clearly briefed about their participation requirements and the time period. Their commitment and consent will be requested if they agree to participate.
4. The experts will be assured of anonymity and confidentiality, with only the research team accessing and retaining their contact details.

#### Questionnaire rounds

##### a) For factors in readiness assessment template and decision making on test and treat options (2-3 rounds):

A comprehensive list of factors relevant for both the readiness assessment as well as decision making for test and treat options was developed through literature review NMPs consultations, and multiple internal team discussions. The factors are categorized into epidemiological, health system, and politico-economic factors (Table 2). For each factor a question that captures that factor as well as a categorization was developed (e.g. for the factor pharmacovigilance the respective suggested question is ‘What is the status of adverse event reporting for any disease in the last 12 months in your country?’ and the suggested categories are ‘a) Adverse Event usually recorded and reported, b) Adverse Event sometimes recorded and reported, c) Adverse Event not recorded or reported, and d) Don't know’) In the first round, the experts will be asked closed-ended questions to seek the importance of each factor for inclusion in the readiness assessment and/or for decision making on different test and treat options.

- The closed-ended options will include: “Important for readiness assessment”, “Important for making decision on different test and treat radical cure options”, “Important for both readiness assessment and decision-making on test and treat options”, and “Not important for readiness assessment or decision-making on test and treat options”
- For each factor, if experts rate the factor as important, then they will be asked if they agree with the question and categorizations we have used to measure the factor.
  - The closed-ended options will include: “yes”, “no”, “don’t know (not my expertise)”, and “need more information to answer”.
- Where possible, an open-ended question will be asked on the optimal threshold criteria for classification of the factor.
- In the second round, the factors and questions/categorizations that did not reach a consensus will be narrowed down. The experts will be made aware of the group results and asked to revisit their responses with regards to those of the group.
- The threshold criteria of the factors will be catergorized according to the feedback from the first round, and the experts will be asked to provide their response on the new categories.
- If needed, additional rounds will be conducted to obtain consensus of the experts on the factors and threshold criteria.

##### b) For recommended combination/s of radical cure test and treat options (2-3 rounds):

- In the first round for this purpose, the 4-5 scenarios developed after literature review, internal team discussions, and feedback from NMP consultations will be presented to the experts.
- The experts will be asked to indicate their recommended radical cure test and treat combinations (i.e. G6PD deficiency screening and radical cure regimen) for each scenario.
- In the second round, agreement on their recommened test and treat combinations will be will be sought for each scenario based on the group results from the first round.
- If needed, further rounds will be conducted to obtain consensus

Table 2. Factors identified for Readiness Assessment:

**a) Epidemiological factors:**

1. Phase of malaria program
2. Vivax caseload
3. G6PDd prevalence
4. G6PDd prevalence heterogeneity
5. Current recommended blood-stage treatment
6. Current recommended liver-stage treatment
7. Efficacy of radical cure regimen

**b) Health system (implementation) factors**

1. Referral system
2. Access
3. Human resource
4. Patient adherence
5. Pharmacovigilance

**c) Politico-ecnomic (enabling) factors**

1. Budget
2. Political will
3. Decision maker's Risk aversion

#### Data collection procedure

- After confirming participation, the online questionnaire link and specific instructions will be emailed to the experts to activate the round 1 for readiness assessment factors.
- Reminder emails will be sent on a weekly basis for 3 weeks if the questionnaire has not been returned.
- Only those expert panel members who return the previous round questionnaire will receive the further round version. These experts will be provided with a feedback about their response and the percentage of participants with relevant response in the last round.
- To maximize retention of experts, the time period between the rounds of Delphi will be kept short to one month [10].
- The above described process will be repeated for scenario validation and radical cure options for each scenario.

#### Data analysis

The response rates of experts will be reported for each round. Descriptive analyses of the socio-demographic details of the experts will be presented. The frequencies and percentages of agreement among the responding experts will be calculated for each item in each round and presented in form of graphs. The criterion for consensus will be kept at ≥75% [11]. For each question, experts who respond with “don’t know (not my expertise)” or “need more information to answer” will be excluded from the analysis. At least 12 experts who have the required expertise to answer definitively will be needed for each question.

Changes in agreement percentage between rounds and any modifications made to the factors and scenarios will also be reported. Fischer’s exact tests will be used to compare the agreement levels between experts of different knowledge domains. A framework described in Table 3 will be used as a guide to proceed with different responses in the Delphi rounds.

Table 3. Framework for handling different responses in the Delphi first round:

| **Response variety** | **Steps to be taken** |
| --- | --- |
| 1. There is 75% or more agreement about the importance of the factor, the suggested question, and the categorization used. | The factor, thw questions and categorization will be included in the readiness assessment template (along with the question and categorization), and/or decision making for test and treat combinations. |
| 1. 75% or more experts in the first round do not regard one factor as important for readiness assessment template and/or for decision making on test and treat combinations | 1. Present the group results to the experts in the second round and ask them to consider the factor one more time.  2. If the consensus is below 75% after the second round, consult with the internal team and NMPs to consider dropping the factor altogether from the readiness assessment and decision tree. |
| 2. There is agreement on the importance of the factor, but less than 75% experts agree with the question we have suggested to measure the factor. | 1. Present the suggested alternative questions along with the original question as options to select from in the second round. Input from NMP consultations will be sought to add additional alternative questions. All additional options as well as the orginal options will be presented and experts asked to select the most suitable question to capture the factor of interest..  2. If the consensus is below 75% after the second round an additional round with the highes ranking options will be conducted until consensus is reached. |
| 3. There is agreement on the importance of the factor and the question used, but less than 75% experts agree with the categorization we have used to capture the factor. | 1. Present the suggested alternative categorizations along with the original categorization as options to select from in the second round.  2. If the consensus is below 75% after the second round an additional round with the highes ranking options will be conducted until consensus is reached |
| 5. Experts have suggested additional factor to be included in the template | 1. If additional factors suggested by experts are also suggested by NMP and if internal team discussion is favourable, the additional factors will be predented to the experts in the second round for consideration.    2. If the consensus is less than 75% after the second round, disregard the additional factor.  3. If the consensus is 75% or more after the second round, consider developing questions and categorizations to measure the factor/s in consultation with the internal team and NMPs. |

### Limitations

The tyranny of experts….
