## Supplementary Table 1 for "Study protocol for development of an options assessment toolkit (OAT) for National Malaria Programs in Asia Pacific to determine best combinations of vivax radical cure for their given contexts"

### Supplementary Table 1. Details of reviewed public health toolkits targeted to health policy makers

| Toolkit | Topic | Audience | Purpose | Method used | Toolkit components | Format | Authors |
| --- | --- | --- | --- | --- | --- | --- | --- |
| 1. WHO Antenatal Care  Recommendations Adaptation Toolkit | Antenatal health | National policymakers | To support countries to systematically adapt the WHO ANC recommendations for country contexts. | 3-step process:  1. Draft toolkit with input from methodologists and regional implementation experts  2. User-testing phase during country stakeholder meetings.   - Stakeholder interviews - Content analysis   3. Toolkit update and development of an instruction manual | 1. Baseline assessment tool  a. Situation analysis tool  b. Recommendation Mapping exercise  c. Country-specific ANC package and SWOT analysis of implementation of new recommendations  2. Qualitative evidence synthesis Slidedoc  Supplementary materials:  a. Implementation considerations  b. Remarks section from each recommendation  c. National ANC guideline template  d. Draft agenda for stakeholder meeting  e. Draft group work materials for stakeholder meeting | Excel sheets, Slides, and report | [Barreix et al 2020](https://doi.org/10.1186/s12961-020-00554-4) |
| 2. MEI Malaria Elimination Toolkit | Malaria | NMCP staff, partner organization, research institutions, evaluators, donors, or others | To build capacity and optimize a country or district’s ability to advance toward elimination. | Not mentioned explicitly | 1. Chemoprevention Options In Advanced Control and Elimination of Malaria (CHOICE) Framework 2. Entomological Surveillance Planning Tool (ESPT) 3. Leadership & Engagement for Improved Accountability & Delivery of Services Framework (LEAD) 4. The District-level Readiness for Elimination of Malaria Tool (DREAM-IT) 5. A Malaria Elimination Guide to Targeted Surveillance and Response in High Risk Populations (HRP) 6. Reactive Case Detection (RACD) Monitoring and Evaluation Tool 7. Malaria Budget Advocacy (MBA) 8. SUSTAIN: A Sustainability and Transition Readiness Assessment Tool for Malaria 9. Primaquine Roll Out Monitoring Pharmacovigilance Tool (PROMPT) | Factsheet, Frameworks, Questionnaires, PDF | [USCF](http://www.shrinkingthemalariamap.org/toolkit) |
| 3. Advocacy Tool Kit | Children and Adolescents Living with and at Risk for HIV | First ladies of African Countries | To advocate for continued uptake of prevention of mother-to-child transmission (PMTCT) of HIV services, increased early infant diagnosis (EID) of HIV, and improved pediatric HIV treatment coverage. | Not mentioned explicitly | 1. Statistics, 2. Key messages, and 3. Key actions that first ladies can undertake | Web-based PDF | [Elizabeth Glaser Pediatric AIDS Foundation (EGPAF) and OAFLA](https://www.pedaids.org/wp-content/uploads/2018/01/OAFLA_20172.pdf) |
| 4. Zero Malaria Starts with Me Toolkit | Advocacy campaign to end malaria in Africa | Governments of African Countries (through Ministries of Health, Finance and Development, and NMPs, the private sector, NGOs, communities, and other members of society. | To enable knowledge sharing and facilitate the adoption of the Zero Malaria Starts with Me movement across the African continent. | Based on tools and materials developed through the Zero Palu! Je m’engage campaign led by the Senegal national Malaria Control Programme (PnLP) and Ministry of Health and Social Action in partnership with Speak Up Africa and PATH. | 1. Quick start guide 2. Module 1: Agenda setting: Tools and guides for defining the goals of malaria advocacy and community engagement in your country as well as research guides for building an evidence base. 3. Module 2: Planning and consultation: Tools and guides for defining objectives, strategy, approaches, actions, and monitoring and evaluation frameworks, as well as for meeting with stakeholders, identifying resources, and consulting with potential partners. 4. Module 3: Political engagement: Guides for launching the campaign, mobilizing political support, and sustaining advocacy. 5. Module 4: Private sector engagement: Guides for launching the campaign with potential private sector partners, building relationships, and fundraising tools. 6. Module 5: Community engagement: Community engagement guides as well as tools for recruiting, supporting, and supervising community malaria champions. 7. Module 6: Making the campaign visible: Contains cross-cutting guidance on working with the media, mobilizing supporters, and engaging via social media. 8. Module 7: Monitoring and evaluation: Contains tools to establish monitoring and evaluation objectives, strategy, and indicators in addition to a guide to programme adjustment based on new developments. | Web-based platform | [African Union Commission with the RBM Partnership to End Malaria](https://endmalaria.org/sites/default/files/Zero%20Malaria%20Toolkit%20Final.pdf) |
| 5. Policy Toolkit for Strengthening Health Sector Reform | Health sector reform | Health sector reform teams and  others involved in making and influencing health policy decisions. | To help health sector reform teams better understand the nature of the political process and develop skills to actively manage that process. | Not mentioned explicitly | 1. Introduction to the Toolkit and the Policy Process 2. Stakeholder analysis guidelines 3. Advocacy guidelines 4. Conflict negotiation guidelines 5. Introduction to strategic management | Web-based PDF and Trainer’s guide | [Latin America and Caribbean Regional Health Sector Reform Initiative](https://www.paho.org/hq/dmdocuments/2010/47-Policy_Toolkit_Strengthening_HSR.pdf) |
| 6. Malaria surveillance, monitoring & evaluation: a reference manual | Malaria surveillance | NMPs, WHO, donors, implementing partners | Adaptability and standardization of malaria surveillance assessment | The Toolkit builds on the PRISM (Performance of Routine Information System Management) model by having a framework based on four objectives that a surveillance assessment can address | 1. Indicator table 2. Protocol outline 3. Desk review guide 4. Data quality assessment guide 5. Question Banks 6. Analysis tools 7. Report outline 8. Assessment evaluation plan 9. Assessment implementation plan | Web-based pdf | [WHO 2018](https://apps.who.int/iris/bitstream/handle/10665/272284/9789241565578-eng.pdf) |
| 7. Malaria Matchbox Tool: An equity assessment tool to improve the effectiveness of malaria programs | Gender and social stratifiers of malaria | National malaria programs together with in-country implementing partners and stakeholders, including civil society and community-based organizations | To support national malaria programs by identifying key affected areas and/or populations and assessing the factors that drive inequities | - Comprehensive revision coordinated by the Global Fund - Several consultations with civil society and community-based organizations | A qualitative analytical framework for the examination of how social, economic, cultural, and gender-related inequities shape malaria and malaria services in a country or region.  The Tool consists of a pre-assessment phase and five detailed modules with step-by-step guidance, as well as illustrative case studies. | Web-based platform | [GF and RBM](https://www.genderhealthhub.org/articles/malaria-matchbox-tool-an-equity-assessment-tool-to-improve-the-effectiveness-of-malaria-programs/) 2019 |
